# The Role of Interleukin-6 in Predicting Adverse Cardiac Outcomes in Asian Hypertrophic Cardiomyopathy

**DOI:** 10.1101/2025.02.23.25322758

**Authors:** Thu-Thao Le, Chee Jian Pua, Tar-Choon Aw, Shiqi Lim, Chengxi Yang, Jennifer Ann Bryant, Yiying Han, Hak Chiaw Tang, Siew Ching Kong, Soon Kieng Phua, Stuart Alexander Cook, Calvin Woon-Loong Chin

## Abstract

**Background:** Interleukin-6 (IL-6) is a pro-inflammatory cytokine implicated in adverse cardiac remodeling and outcomes in various cardiovascular diseases. Its role in hypertrophic cardiomyopathy (HCM), particularly in risk stratification and prognosis, remains underexplored. This study evaluates serum IL-6 levels alongside cardiac magnetic resonance (CMR) imaging parameters as predictors of major adverse cardiac events (MACE) in patients with apical HCM (ApHCM) and non-apical HCM (Non-ApHCM).

**Method:** A cohort of 255 patients was recruited between 2013 and 2023, with blood samples collected for genetic analysis and biomarker testing. A subset of 196 patients underwent CMR imaging to assess left ventricular morphology, function, and myocardial fibrosis. MACE was defined as the composite occurrence of ventricular arrhythmias, heart failure, atrial fibrillation, stroke, or all-cause mortality.

**Results:** Patients with elevated IL-6 levels (above the median) demonstrated significantly higher MACE rates compared to those with lower levels (7.5 vs. 3.3 events/100 patient-years; pairwise log-rank P < 0.001). IL-6 levels correlated with adverse CMR markers, including myocardial fibrosis, increased left atrial volume, and impaired strain, regardless of HCM subtype. Combining IL-6 levels with morphological classification revealed stepwise risk stratification, with MACE rates escalating from 0.9 (ApHCM with low IL-6) to 8.8 events/100 patient-years (Non-ApHCM with high IL-6) (pairwise log-rank P < 0.001).

**Conclusion:** This study highlights IL-6 as a critical biomarker associated with adverse outcomes in HCM. When integrated with morphological assessment, IL-6 enhances risk stratification and provides a novel approach to assessing disease severity. These findings underscore the potential for IL-6 to guide clinical decision-making and inform therapeutic strategies in HCM.

## INTRODUCTION

Hypertrophic cardiomyopathy (HCM) is a genetically diverse cardiac disorder with a wide spectrum of clinical presentations and disease progression, requiring comprehensive risk stratification to optimize patient management^1^. Apical hypertrophic cardiomyopathy (ApHCM), a morphological variant of hypertrophic cardiomyopathy (HCM), accounts for up to 40% of HCM cases in Asian populations^2–4^. This variant follows a different clinical trajectory, typically presenting a lower risk of heart failure (HF) and a more favourable prognosis compared to non-apical hypertrophic cardiomyopathy (Non-ApHCM)^5–10^.

Risk stratification in HCM traditionally relies on biomarkers such as N-terminal pro-B-type natriuretic peptide (NT-proBNP) and high-sensitivity troponin T (hsTnT), alongside key imaging parameters like myocardial fibrosis and left ventricular strain^11–16^. While these markers are well-established, their prognostic value across HCM subtypes has not been fully elucidated^1,17^. Understanding how these indicators differ between ApHCM and Non-ApHCM could provide critical insights for tailoring clinical management strategies.

Emerging evidence suggests that inflammatory processes play a significant role in the pathophysiology of HCM, contributing to adverse remodeling and disease progression^18^. Among inflammatory markers, interleukin-6 (IL-6) has been increasingly associated with cardiac hypertrophy, fibrosis, and functional impairment in cardiovascular diseases^19,20^. Elevated IL-6 levels have been reported in HCM, yet their specific prognostic significance, particularly in predicting adverse cardiac events, remains underexplored.

This study aims to address this gap by investigating the prognostic role of IL-6 in HCM, with a particular focus on its utility as a biomarker for adverse cardiac outcomes across ApHCM and Non-ApHCM subtypes. By integrating IL-6 levels with cardiac magnetic resonance (CMR) imaging parameters, this study seeks to provide novel insights into the inflammatory mechanisms underlying HCM progression and enhance risk stratification for optimized clinical care.

## METHODS

### Study population

A total of 255 unrelated patients with a clinical diagnosis of ApHCM or other HCM morphologies (Non-ApHCM) based on contemporary guidelines were prospectively recruited from the Cardiomyopathy Clinic at the National Heart Centre Singapore from 2013 and followed up until December 2023^21,22^. All participants provided written informed consent. The study was conducted in accordance with the Local Tissue Acts and the Declaration of Helsinki and approved by the SingHealth Centralized Institutional Review Board.

### Sequencing and variant classification

Genetic evaluation of variation in established HCM-associated genes was performed using Illumina TruSight Cardio panel in all HCM patients as previously described^23^. Briefly, libraries were individually indexed, purified and enriched for genes related to HCM. Pooled libraries were sequenced using Illumina MiSeq (v2 kit), MiniSeq (Mid Output kit) or NextSeq 500 (Mid Output v2 kit) benchtop sequencers using paired– end, 150bp reads. Patients were categorised under four genetic strata. Understudied population including Singaporean HCM cases had previously found to have a greater excess of variant of unknown significance compared to the White population^24^. Overall burden of rare protein-altering variants in HCM cases compared to controls was comparable between the Singaporean and White populations. Patients carrying at least at least one putative causative rare variant (maximum population allele frequency <0.00004) in HCM-related genes or local founder/recurrent variants (TNNT2:p.Arg286His and TNNI3:p.Arg79Cys) were considered as genotype-positive (G+). Patients with G+ were sub-categorised into (i) those carrying variants in thick filament sarcomeric genes (*MYBPC3*, *MYH7*, *MYL2, MYL3*), (ii) thin filament sarcomeric genes (*ACTC1*, *TNNC1*, *TNNI3*, *TNNT2*, *TPM1*), and (iii) other genotypes including genocopies (*CSRP3*, *FHL1*, *GLA*, *PLN*, *PRKAG2*, *LAMP2*). All other patients were classified as genotype-negative (G-).

### Variant curation pipeline

Raw sequencing data were demultiplexed, trimmed and mapped to UCSC GRCh37/hg19 reference genome before variant calling using GATKv3.8.1 HaplotypeCaller^25^. All variants were annotated using Ensembl Variant Effect Predictor (VEP v102)^26^ with plugins from dbNSFP (version 4.1a)^27^, gnomAD (version r2.1.1)^28^, ClinVar (version 20210213)^29^ and LOFTEE^30^. Variant classification was subsequently performed according to the ACMG/AMP guidelines^31^.

### Serum biomarkers

Blood samples were collected and stored at −80°C. Serum NT-proBNP, hsTnT, C-reactive protein (CRP) and IL-6 were assayed using electrochemiluminescence immunoassay (NT-proBNP, hsTnT, and IL-6) or particle-enhanced immunoturbidimetric assay (CRP) on the Cobas C701 analyzer (Roche Diagnostics Asia-Pacific, Singapore).

The manufacturer-reported lower limit of detection (LOD) was 5 pg/mL for NT-proBNP, 3 pg/mL for hsTnT, 0.3 mg/L for CRP, and 1.5 pg/mL for IL-6.

Concentrations lower than the detection levels in the participants were assigned a value equivalent to half of LOD.

### Cardiovascular magnetic resonance imaging

A subset of participants (n = 196) underwent CMR (Siemens Aera 1.5T, Erlangen, Germany or Philips Ingenia 3T, Best, The Netherlands). Balanced steady-state free precession cine images of the long-axis views and short-axis covering the heart from base to the apex were acquired (1.6 x 1.3 x 8 mm^3^, 30 phases per cardiac cycle). Late gadolinium enhancement (LGE) as well as native and postcontrast myocardial T1 mapping (Modified Look-Locker Inversion-recovery sequence, Siemens Aera 1.5T) were obtained with 0.1 mmol/kg of gadobutrol (Gadovist, Bayer Pharma AG, Germany).

Deidentified imaging data were analyzed at the National Heart Research Institute Singapore (NHRIS) CMR Core Laboratory using CVI42 (Circle Cardiovascular Imaging, Calgary, Canada) by individuals who were blinded to clinical, genotyping and outcome data. Cardiac volumes, left ventricular (LV) mass and maximal wall thickness (WT_max_) were measured using standardized protocols and indexed to body surface area as previously published^32,33^. Remodeling index (RI), a measure of advanced hypertrophy derived from wall-stress model, was calculated from the LV end-diastolic volume and WT_max_^34^. LV myocardial strains were measured from short-axis, 4-chamber, 3-chamber and 2-chamber long-axis cine images^35^. Myocardial replacement fibrosis was assessed using LGE images, while extracellular volume (ECV) and interstitial fibrosis volume, measures of myocardial interstitial fibrosis, were quantified using T1 maps^36,37^.

### Outcome definitions

The primary outcome of major adverse cardiac event (MACE) was defined as a composite of first occurrence of ventricular arrhythmic, heart failure, atrial fibrillation, stroke, and all-cause mortality, according to previously published study^15^. MACE was adjudicated by an independent cardiologist who reviewed the electronic medical records. The event rate was presented as number of events per 100 person-years.

### Statistical analysis

Categorical variables were presented as frequency (percentage) and continuous variables were presented as mean±standard deviation or median (interquartile range), depending on normality. The Shapiro-Wilk test was used to assess the distribution of continuous variables. Differences in characteristics between the groups were analyzed by the parametric Student t test and 1-way ANOVA, or the nonparametric Mann-Whitney U test and Kruskal-Wallis test for continuous data, depending on the normality of the data. Categorical data were compared using the χ^2^ test.

Linear regression was used to assess the associations between morphological and biochemical predictors of outcomes and CMR measures, adjusted for age, sex, metabolic comorbidities and genotype.

Univariable Cox-proportional hazards models were used to examine the prognostic importance of morphologies (ApHCM and Non-ApHCM) in HCM and its subgroups, categorized by study means or medians, depending on the normality of the data. The proportional hazards assumption was evaluated by log-minus-log plot.

MACE-free survival of groups stratified using HCM morphologies, CMR features and biomarkers were examined using the Kaplan-Meier method and compared with log-rank test.

All statistical analyses were performed using SPSS, version 28 (IBM, Inc, Armonk, NY) and GraphPad Prism, version 10.2.0 (GraphPad Software, San Diego, CA). A 2-sided P<0.05 was considered as statistically significant.

## RESULTS

### Clinical characteristics and genetics

Among the 255 HCM patients recruited, 71 (27.8%) had ApHCM, with a significantly higher proportion of males compared to Non-ApHCM (90.1% versus 69.6%, P<0.001) (Table 1). Most patients, regardless of morphologies, had no to mild symptoms, with the majority classified as New York Heart Association (NYHA) functional class I-II. ApHCM patients had significantly higher proportions of metabolic risk factors compared to Non-ApHCM (23.9% versus 9.8% with diabetes mellitus, P=0.003; and 60.6% versus 37.5% with hypertension, P<0.001), but a lower proportion with rare protein-altering variants (25.4% versus 49.5%, P<0.001).

**Table 1.**
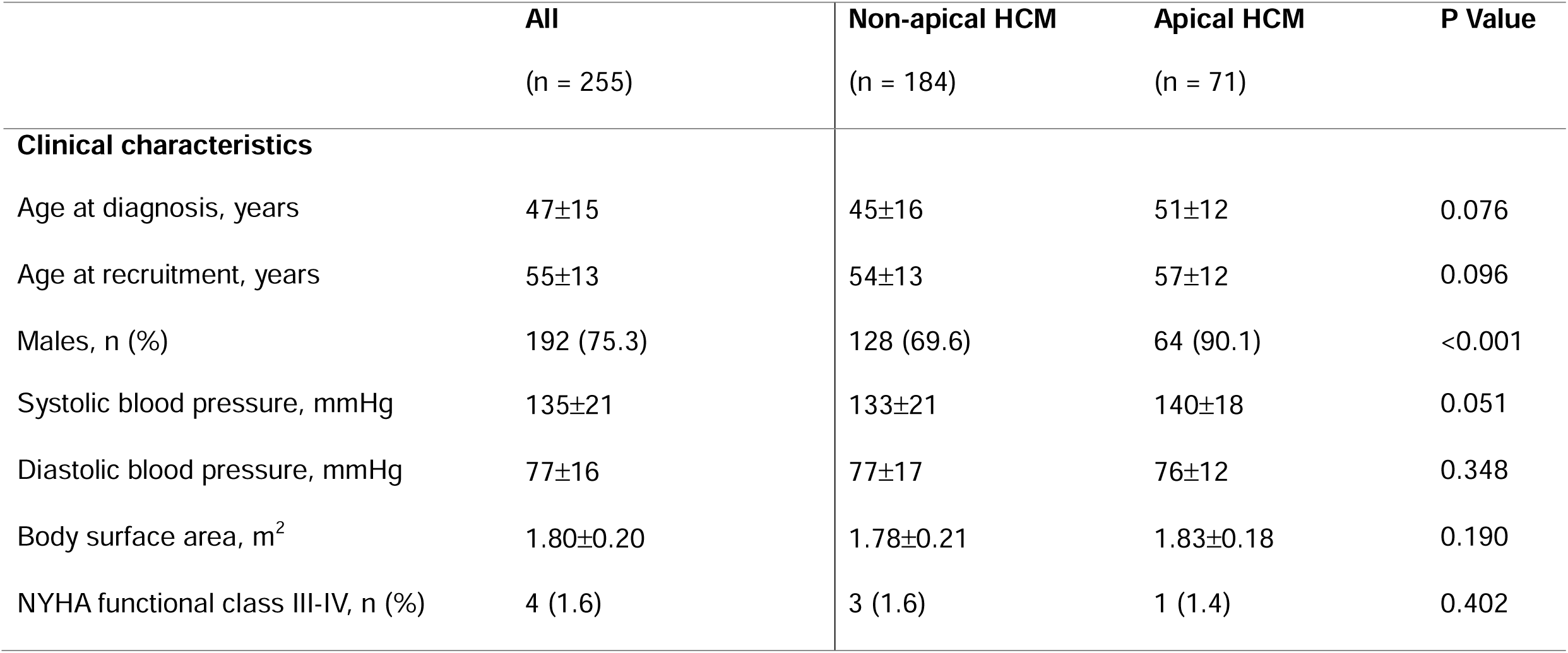
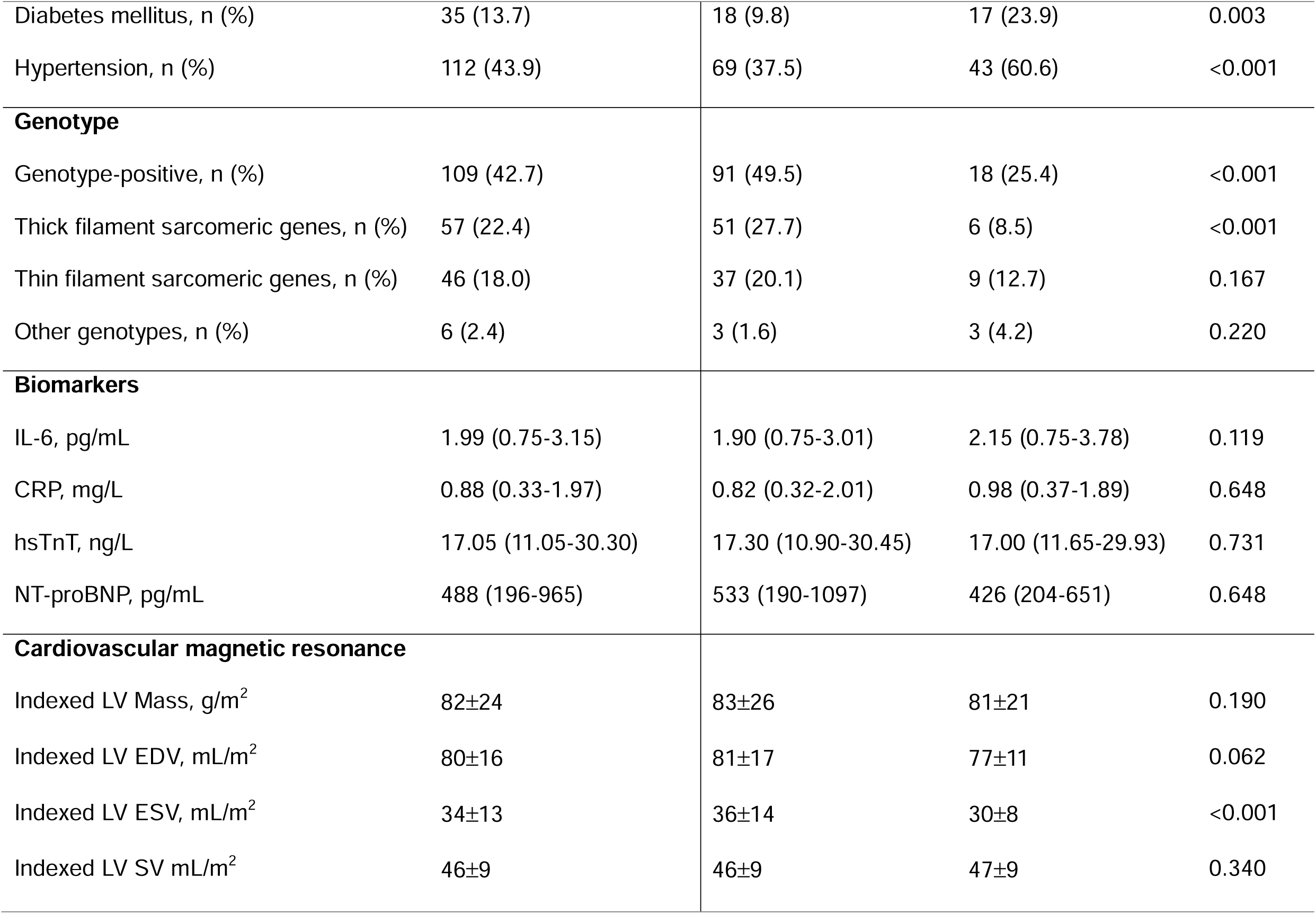
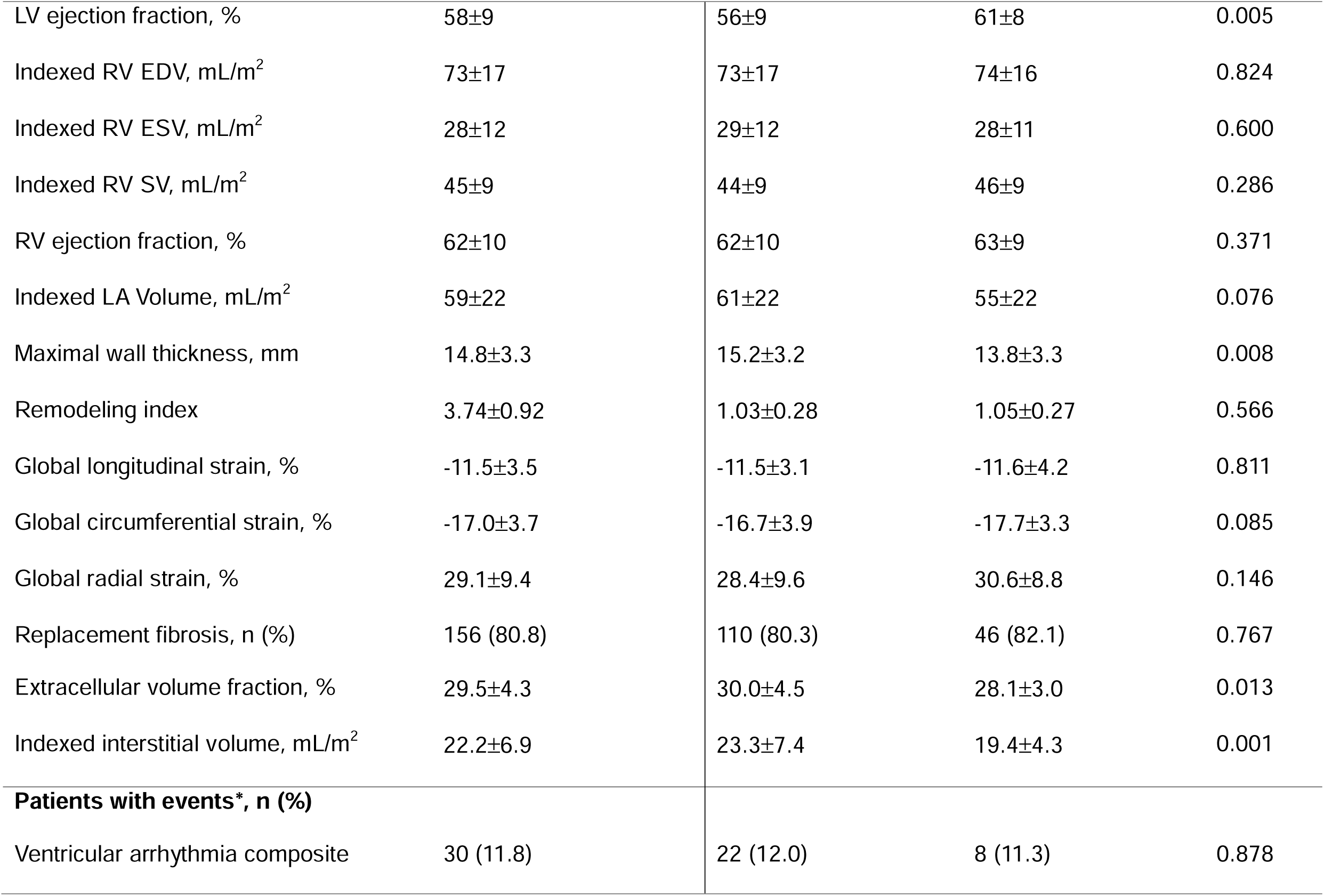
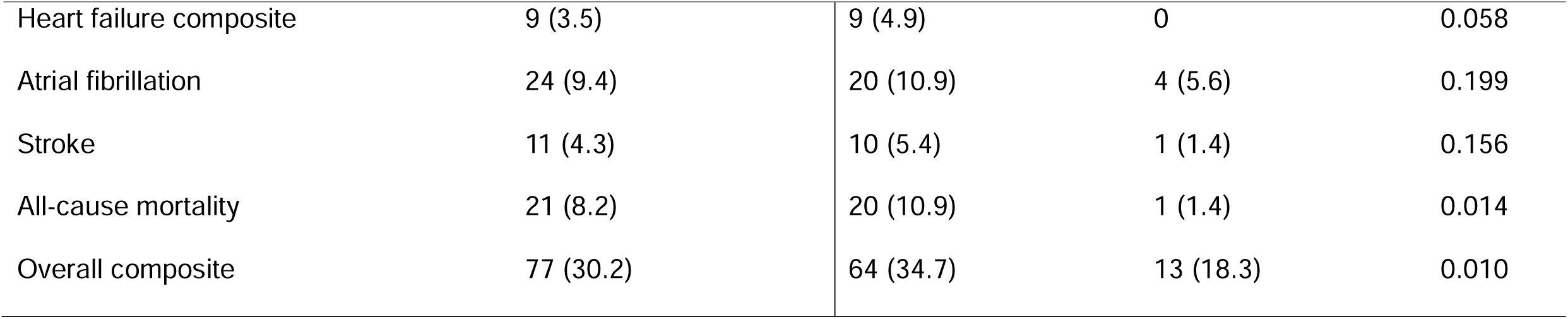
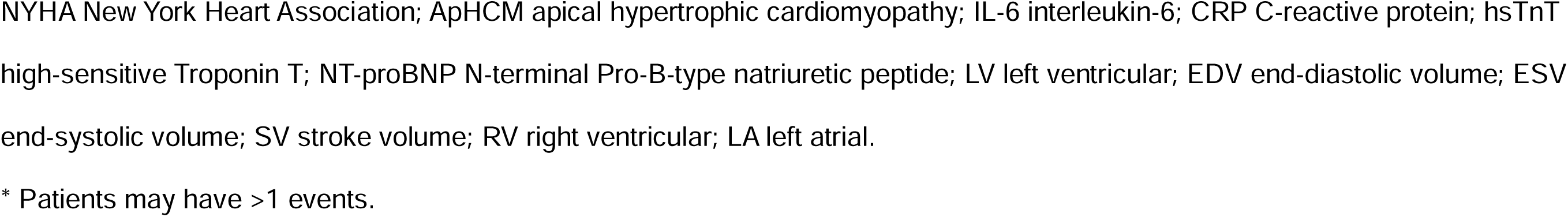
Clinical characteristics, genotypes, circulating biomarkers, and cardiovascular magnetic resonance imaging parameters at baseline and clinical outcomes. Comparison of clinical characteristics and cardiovascular magnetic resonance imaging parameters between Low-risk (apical hypertrophic cardiomyopathy with Interleukin-6 levels below median), Moderate-risk (apical hypertrophic cardiomyopathy with Interleukin-6 levels above median, or non-apical hypertrophic cardiomyopathy with Interleukin-6 levels below median), and High-risk (non-apical hypertrophic cardiomyopathy with Interleukin-6 levels above median).

### Circulating biomarkers

The majority of HCM patients had elevated IL-6, hsTnT and NT-proBNP values (IL-6: 1.99 (0.75 – 3.15) pg/mL; hsTnT: 17.05 (11.05 – 30.30) ng/L; and NT-proBNP: 488 (196 – 965) pg/mL), while CRP remained normal (0.88 (0.33 – 1.97) mg/L), per Roche Diagnostics reference ranges. All 4 biomarkers showed similar values between ApHCM and Non-ApHCM (Table 1). When adjusting for age, sex, hypertension, and diabetes, ApHCM was associated with increased CRP (OR: 2.42 [95%CI 1.61 – 3.29], P<0.001), IL-6 (OR: 2.60 [95%CI 1.96 – 3.44], P<0.001) and hsTnT (OR: 1.46 [95%CI 1.13 – 1.89], P=0.003).

### Cardiovascular magnetic resonance imaging parameters

Non-ApHCM patients had significantly worse LV ejection fraction (56±9% versus 61±8%, P=0.005) and thicker myocardial wall (15.2±3.2 mm versus 13.8±3.3 mm, P=0.008) compared to ApHCM (Table 1). While the proportions of patients with replacement fibrosis were similar, Non-ApHCM patients exhibited increased interstitial fibrosis compared to those with ApHCM (ECV: 30.0±4.5% versus 28.1±3.0, P=0.013; indexed interstitial volume: 23.3±7.4 g/m^2^ versus 19.4±4.3 g/m^2^, P=0.001).

Among the circulating biomarkers, only NT-proBNP was associated with CMR measures of adverse LV remodeling and myocardial fibrosis. hsTnT was only associated with increased replacement fibrosis, while no associations were found between any CMR parameters and CRP or IL-6 (Online Supplemental - Table S1).

### Prognostic Value of IL-6 in Predicting HCM Outcomes

Over a follow-up period of 6.7±2.4 years, 77 patients experienced MACEs (Table 1). Patients with elevated IL-6 levels had significantly worse cardiac outcomes, with event rates of 7.5 versus 3.3 events per 100 patient-years for those above versus below the median IL-6 levels (pairwise log-rank P < 0.001). Elevated IL-6 was independently associated with a twofold higher risk of MACEs (HR: 2.428, 95% CI: 1.483–3.975, P < 0.001), regardless of HCM morphology (Figures 1 and 2).

**Figure 1.** Hazard ratios predicting major adverse cardiac events. Subgroup analysis of the association between apical hypertrophic cardiomyopathy (ApHCM) and non-apical hypertrophic cardiomyopathy (Non-ApHCM) and major adverse cardiac events.

**Figure 2.**
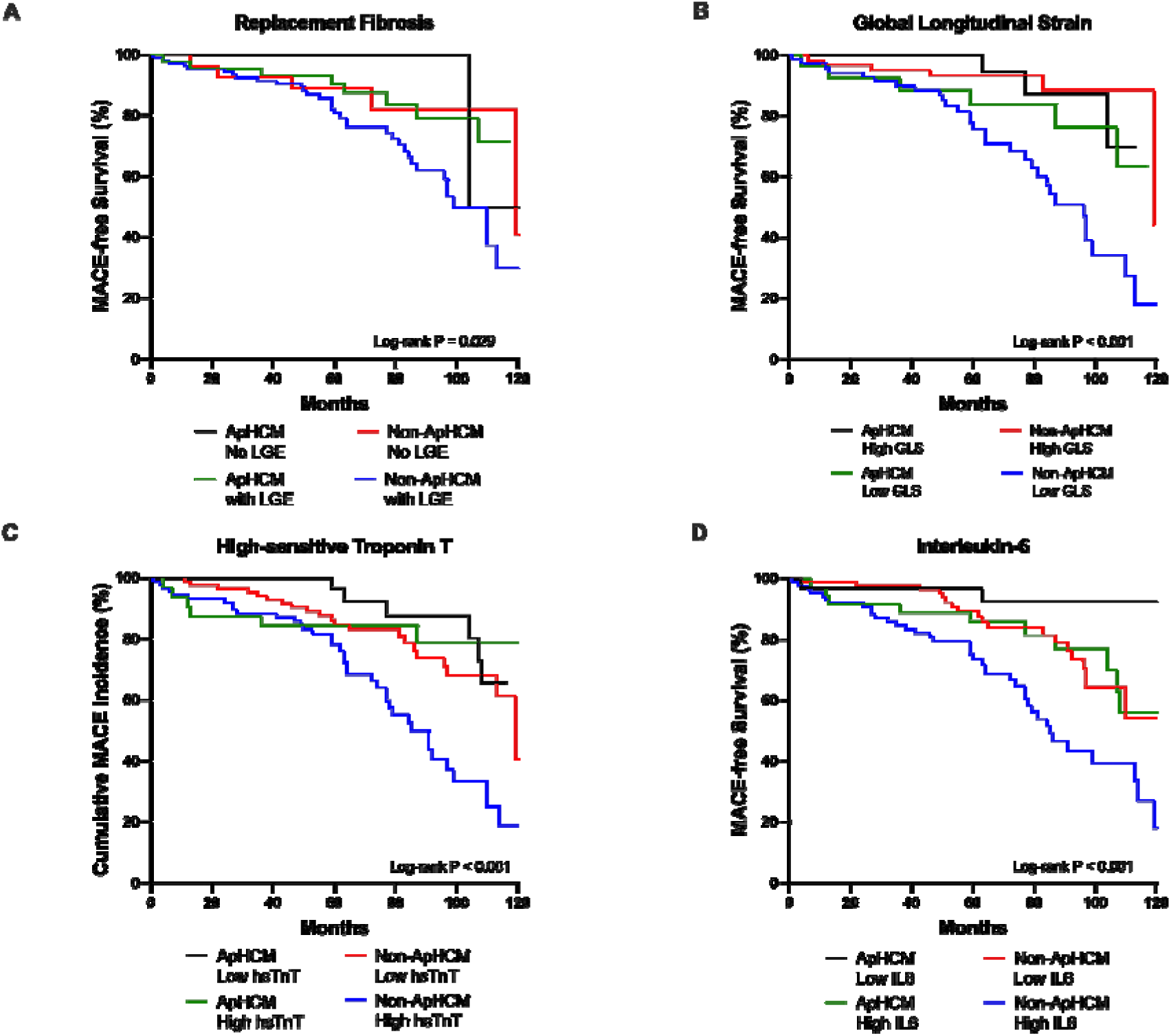
Major adverse cardiac event-free survival in hypertrophic cardiomyopathy patients. Survival curves for major adverse cardiac events among apical hypertrophic cardiomyopathy and non-apical hypertrophic cardiomyopathy, with subgroup risk-stratification by (**A**) presence of myocardial fibrosis, (**B**) global longitudinal strain below and above study mean, (**C**) high-sensitive troponin T below and above median levels, and (**D**) interleukin-6 below and above median levels.

When stratified by both IL-6 levels and morphological subtypes, Non-ApHCM patients with elevated IL-6 levels had the highest risk of MACEs (8.8 events per 100 patient-years), followed by ApHCM patients with high IL-6 (4.2 events per 100 patient-years), Non-ApHCM with low IL-6 (3.7 events per 100 patient-years), and ApHCM with low IL-6 (0.9 events per 100 patient-years) (pairwise log-rank P < 0.001). These findings demonstrate the additive prognostic value of combining inflammatory biomarker levels with morphological subtypes.

ApHCM patients, despite a higher burden of metabolic comorbidities, exhibited lower event rates compared to Non-ApHCM patients, particularly when IL-6 levels were elevated. Additionally, Non-ApHCM patients with high IL-6 levels showed significantly greater interstitial fibrosis burden (Supplemental Table S2), as reflected by indexed interstitial volume on cardiac magnetic resonance imaging.

## DISCUSSION

This study demonstrates for the first time that elevated IL-6 levels are independently associated with an increased risk of MACEs in patients with HCM, offering novel insights into the inflammatory underpinnings of disease progression. By integrating IL-6 levels with morphological subtypes, we offer a significant advancement in risk stratification for adverse outcomes, emphasizing the importance of inflammation in the pathophysiology of HCM.

### The Role of IL-6 in HCM Pathology

Hypertension and diabetes are common in older HCM patients^1,5,38,39^ and are linked to a greater burden of myocardial fibrosis, impaired diastolic function, and elevated biomarkers such as NT-proBNP and troponin levels^1,40^. Our observation of significantly higher incidence of hypertension and diabetes in ApHCM patients aligns with findings from other cohorts and likely reflects the accumulation of metabolic comorbidities with age, as ApHCM patients in this study were generally older^41,42^. Even after adjusting for age, sex, and metabolic comorbidities, ApHCM patients exhibited significantly higher levels of CRP, IL-6 and hsTnT compared to Non-ApHCM, suggesting a more pronounced inflammatory and myocardial injury profile. Chronic low-grade inflammation, characterized by increased levels of inflammatory cytokines such as CRP and IL-6, is well-documented in HCM patients^19^. While CRP has been widely studied as a marker of inflammation in cardiovascular diseases, its utility in HCM appears to be limited compared to IL-6. CRP primarily reflects systemic inflammation and is less specific in capturing the localized inflammatory processes within the myocardium that drive HCM progression^43^. On the other hand, experimental studies have demonstrated that IL-6 directly contributes to adverse cardiac remodeling, serving as a key driver of LV hypertrophy, myocardial fibrosis, and dysfunction^44–47^. These effects are thought to arise from IL-6’s ability to activate fibroblasts, promote extracellular matrix deposition, and stimulate hypertrophic signaling pathways within cardiomyocytes. Furthermore, IL-6 is elevated in HCM patients with moderate to severe histological fibrosis, reinforcing its association with the underlying pathology of the disease^19^.

Importantly, while IL-6 levels were positively correlated with the extent of replacement fibrosis as assessed by LGE images, no significant correlation was found between IL-6 and LV systolic function or interstitial fibrosis^48,49^. This suggests that IL-6 may exert its effects through complex, multifactorial pathways rather than a straightforward, linear relationship with imaging-based metrics of fibrosis or ventricular function. These pathways could involve interactions with other cytokines, cellular signaling mechanisms, and immune-mediated processes, highlighting the unique and intricate role of IL-6 in HCM progression.

### Combining IL-6 and HCM Morphology for Enhanced Risk Stratification

Despite the lack of evidence supporting IL-6 as a diagnostic tool for identifying patients at risk of worsening cardiac function and myocardial fibrosis, our findings highlight its prognostic value for predicting adverse cardiac outcomes in HCM patients. This aligns with recent studies showing that IL-6 has a stronger association with adverse cardiac outcomes than CRP in other cardiac conditions^50–52^.

Both HCM morphology and IL-6 levels were independently strong predictors of prognosis, yet they likely reflect distinct underlying myocardial pathologies stemming from different biological mechanisms. HCM morphology provides insight into structural and genetic determinants of the disease, while IL-6 levels capture the inflammatory burden that contributes to pathological remodeling. By stratifying IL-6 levels, we could delineate clinically meaningful differences in inflammatory risk, highlighting the complex and multifaceted role of IL-6 in HCM pathophysiology and the disease’s inherent heterogeneity.

The combination of HCM morphology and IL-6 levels provided enhanced risk stratification, offering novel insights that extended beyond the predictive capabilities of either factor alone. Specifically, patients with the Non-ApHCM variant and elevated IL-6 were identified as the highest-risk subgroup for MACEs. This finding is particularly noteworthy because, despite having fewer metabolic risk factors compared to ApHCM patients with similarly elevated IL-6 levels, Non-ApHCM patients exhibited MACE outcomes comparable to the highest-risk groups identified using conventional CMR markers or hsTnT.

Interestingly, genetic analysis revealed that approximately half of the Non-ApHCM patients carried pathogenic/likely pathogenic HCM genes, predominantly involving sarcomeric proteins. These genetic abnormalities are closely linked to increased myocardial fibrosis^1^, and when compounded by chronic inflammation, as indicated by elevated IL-6 levels, they likely drive the heightened fibrosis burden in this subgroup. This is reflected in a significantly higher indexed interstitial volume, a surrogate marker for absolute interstitial fibrosis^37^. In contrast, ApHCM patients had a lower prevalence of sarcomeric mutations, and those with elevated IL-6 levels appeared to represent a relatively low-risk group, likely to respond well to standard treatment regimens without the need for additional interventions.

These findings suggest that Non-ApHCM patients with elevated IL-6 patients may benefit from more aggressive management strategies, including novel anti-inflammatory therapies, aimed at mitigating chronic inflammation’s contribution to disease progression. Conversely, ApHCM patients with elevated IL-6, given their lower overall risk, may not require such intensive approaches. This tailored treatment approach highlights the clinical utility of integrating IL-6 measurements with morphological classification in HCM management.

While genetic testing and imaging markers such as myocardial fibrosis, LV hypertrophy and LA size remain cornerstones of HCM risk assessment and management guidelines^53^, our study underscores the added value of incorporating inflammatory biomarkers like IL-6. Among patients with similar adverse cardiac remodeling observed on CMR imaging, Non-ApHCM morphology was associated with significantly higher MACE risk, likely due to the interplay between genetic predisposition and inflammation. Notably, in patients without significant cardiac remodeling, prognosis was comparable across morphological subtypes, further emphasizing the complex interaction of structural, genetic, and inflammatory factors in shaping clinical outcomes.

The integration of IL-6 with HCM morphology in risk stratification represents a novel and promising approach that enhances our ability to identify high-risk subgroups and inform targeted therapeutic strategies. This combined framework moves beyond traditional imaging and genetic assessments, providing a more comprehensive and individualized evaluation of risk in HCM patients.

### Study limitation

Given the observational nature of this study, we did not explore causality between factors such as genetic variants, inflammatory pathways, and myocardial fibrosis associated with specific HCM morphologies and circulating IL-6 levels. Subgroup analyses relied on study mean or median values due to the absence of established reference ranges for interstitial fibrosis and biomarkers within this population. While this categorical approach offers insights into how different parameters interact with HCM morphologies, it is limited by the small sample size and low event rates, particularly for ApHCM variant. Consequently, further validation in larger cohorts is necessary to confirm these findings and refine risk stratification models for more robust prognostic evaluation.

## CONCLUSION

This study establishes elevated IL-6 levels as an independent inflammatory predictor of adverse cardiac outcomes in HCM, irrespective of morphologic subtype. While ApHCM patients demonstrated a generally better prognosis for MACEs compared to Non-ApHCM patients, the inclusion of IL-6 in risk stratification models significantly enhanced their predictive accuracy. These results advocate for integrating IL-6 as part of a multifactorial approach to risk stratification in HCM management. Further studies with larger cohorts are essential to validate and refine these models and explore the therapeutic implications of targeting inflammation in HCM.

## Data Availability

All data produced in the present study are available upon reasonable request to the authors.

## ABBREVIATION

HCM: Hypertrophic cardiomyopathy
ApHCM: Apical hypertrophic cardiomyopathy
Non-ApHCM: Non-apical hypertrophic cardiomyopathy
CMR: Cardiovascular magnetic resonance
NT-proBNP: N-terminal pro-B-type natriuretic peptide
hsTnT: High-sensitive troponin T
CRP: C-reactive protein
IL-6: Interleukin-6
LOD: Lower limit of detection
LGE: Late-gadolinium enhancement
LV: Left ventricular/ventricle
LA: Left atrial/atrium
RI: Remodeling index
WT_max_: Maximal wall thickness
ECV: Extracellular volume fraction
MACE: Major adverse cardiac event
NYHA: New York Heart Association
OR: Odd ratio
HR: Hazard ratio
CI: Confidence interval

## Acknowledgements

We thank the radiographers at the National Heart Centre Singapore for their assistance in the study.

