## Supplementary material for "The Role of Interleukin-6 in Predicting Adverse Cardiac Outcomes in Asian Hypertrophic Cardiomyopathy": Online Supplemental

**Table S1. Associations between circulating biomarkers and cardiovascular magnetic resonance parameters**

|  | CRP | IL6 | hsTnT | NT-prBNP |
| --- | --- | --- | --- | --- |
| Indexed LV EDV | 0.98 [0.96-1.01] | 0.99 [0.97-1.03] | 1.00 [0.99-1.02] | 1.01 [1.00-1.02] ^§^ |
| LV ejection fraction | 0.98 [0.91-1.05] | 1.02 [0.97-1.06] | 0.97 [0.94-1.00] | 0.97 [0.96-0.98] ^§^ |
| Indexed LV mass | 0.99 [0.97-1.01] | 0.99 [0.97-1.02] | 1.01 [1.00-1.02] | 1.02 [1.01-1.02] ^§^ |
| Wall thickness | 0.92 [0.83-1.03] | 0.93 [0.80-1.09] | 1.03 [0.94-1.13] | 1.10 [1.04-1.16] ^§^ |
| Indexed LA Volume | 0.99 [0.97-1.01] | 0.99 [0.97-1.01] | 1.00 [0.99-1.02] | 1.02 [1.01-1.02] ^§^ |
| Global longitudinal strain | 0.94 [0.80-1.10] | 1.12 [0.99-1.25] | 1.08 [0.99-1.19] | 1.17 [1.12-1.21] ^§^ |
| Replacement fibrosis | 1.31 [0.25-4.89] | 2.18 [0.56-6.52] | 2.51 [1.31-4.47] ^§^ | 2.70 [1.78-3.97] ^§^ |
| Extracellular volume | 0.98 [0.80-1.26] | 1.03 [0.83-1.32] | 1.02 [0.91-1.17] | 1.07 [1.02-1.12] ^§^ |
| Indexed interstitial volume | 0.96 [0.91-1.03] | 0.98 [0.90-1.11] | 0.99 [0.95-1.05] | 1.07 [1.03-1.09] ^§^ |

LV left ventricular; EDV end-diastolic volume; LA left atrial; CRP C-reactive protein; IL6 interleukin-6; hsTnT high-sensitive Troponin T; NT-proBNP N-terminal pro-B-type natriuretic peptide. Values presented as odd ratio [95% confidence interval], adjusted for age, sex, hypertension and diabetes.

**Table S2. Clinical characteristics, genotype, circulating biomarkers, cardiovascular magnetic resonance parameters and outcomes among groups stratified by both hypertrophic cardiomyopathy morphologies and interleukin-6 levels.**

|  | Interleukin-6 levels  Below Median | | Interleukin-6 levels  Above Median | | P value |
| --- | --- | --- | --- | --- | --- |
|  | **ApHCM**  **(n = 35)** | **Non-ApHCM**  **(n = 86)** | **ApHCM**  **(n = 38)** | **Non-ApHCM**  **(n = 78)** |  |
| Clinical characteristics |  |  |  |  |  |
| Age at diagnosis, years | 47±12 | 42±15 | 53±12 | 48±15 | 0.001 |
| Age at recruitment, years | 54±11 | 52±13 | 60±12 | 57±13 | 0.004 |
| Males, n (%) | 32 (91.4) | 64 (74.4) | 32 (84.2) | 64 (82.1) | <0.001 |
| Systolic blood pressure, mmHg | 138±17 | 129±18 | 143±20 | 137±24 | 0.002 |
| Diastolic blood pressure, mmHg | 75±10 | 74±13 | 77±13 | 81±20 | 0.035 |
| Body surface area, m^2^ | 1.85±0.16 | 1.75±0.19 | 1.83±0.18 | 1.82±0.23 | 0.045 |
| NYHA functional class III-IV, n (%) | 1 (2.9) | 2 (2.3) | 0 (0) | 1 (1.3) | 0.896 |
| Diabetes mellitus, n (%) | 7 (20.0) | 7 (8.1) | 11 (28.9) | 10 (12.8) | <0.001 |
| Hypertension, n (%) | 15 (42.9) | 33 (38.4) | 28 (73.7) | 36 (46.2) | <0.001 |
| Genotype |  |  |  |  |  |
| Genotype-positive, n (%) | 10 (28.6) | 50 (58.1) | 8 (21.1) | 41 (52.6) | 0.005 |
| Thick filament sarcomeric genes, n (%) | 2 (5.7) | 26 (30.2) | 4 (10.5) | 25 (32.1) | 0.002 |
| Thin filament sarcomeric genes, n (%) | 6 (17.1) | 20 (23.3) | 3 (7.9) | 16 (20.5) | 0.241 |
| Other genotypes, n (%) | 2 (5.7) | 3 (3.5) | 1 (2.6) | 0 (0) | 0.275 |
| Biomarkers |  |  |  |  |  |
| CRP, mg/L | 0.51 (0.15-1.07) | 0.59 (0.15-1.16) | 1.32 (0.71-2.80) | 1.35 (0.49-3.32) | <0.001 |
| hsTnT, ng/L | 14.15 (9.82-32.55) | 16.30 (9.87-26.60) | 20.65 (15.18-30.50) | 18.80 (13.23-35.20) | 0.016 |
| NT-proBNP, pg/mL | 280 (147-592) | 508 (163-961) | 486 (250-1022) | 567 (238-1435) | 0.051 |
| Cardiovascular magnetic resonance | |  |  |  |  |
| Indexed LV Mass, g/m^2^ | 80±20 | 81±28 | 81±22 | 84±23 | 0.794 |
| Indexed LV EDV, mL/m^2^ | 82±10 | 82±15 | 74±11 | 81±19 | 0.093 |
| Indexed LV ESV, mL/m^2^ | 31±8 | 34±11 | 29±8 | 37±17 | 0.030 |
| Indexed LV SV mL/m^2^ | 50±9 | 48±10 | 44±9 | 43±8 | 0.003 |
| LV ejection fraction, % | 62±8 | 58±9 | 60±8 | 55±9 | 0.005 |
| Indexed RV EDV, mL/m^2^ | 80±18 | 75±19 | 68±11 | 71±15 | 0.015 |
| Indexed RV ESV, mL/m^2^ | 30±13 | 29±13 | 25±8 | 28±12 | 0.431 |
| Indexed RV SV, mL/m^2^ | 50±8 | 46±10 | 42±8 | 42±8 | <0.001 |
| RV ejection fraction, % | 64±9 | 63±10 | 63±9 | 61±10 | 0.063 |
| Indexed LA Volume, mL/m^2^ | 50±14 | 60±20 | 59±27 | 62±25 | 0.128 |
| Maximal wall thickness, mm | 13.2±2.8 | 15.0±3.3 | 14.3±3.7 | 15.4±3.0 | 0.023 |
| Remodeling index | 4.18±0.87 | 3.64±0.83 | 3.82±1.06 | 3.56±0.89 | 0.018 |
| Global longitudinal strain, % | -12.9±3.6 | -12.1±3.1 | -10.4±4.4 | -10.9±3.1 | 0.010 |
| Global circumferential strain, % | -18.8±2.9 | -16.9±3.5 | -16.7±3.4 | -16.3±4.3 | 0.038 |
| Global radial strain, % | 33.5±8.0 | 28.9±9.0 | 28.0±8.8 | 27.9±10.3 | 0.062 |
| Replacement fibrosis, n (%) | 20 (57.1) | 58 (67.4) | 26 (68.4) | 53 (67.9) | 0.447 |
| Extracellular volume fraction, % | 27.5±3.6 | 29.9±4.0 | 28.7±2.1 | 30.2±5.2 | 0.169 |
| Indexed interstitial volume, mL/m^2^ | 19.6±5.1 | 21.6±5.8 | 19.0±3.3 | 25.1±8.6 | 0.007 |
| Patients with events*, n (%) |  |  |  |  |  |
| Ventricular arrhythmia composite | 2 (5.7) | 10 (11.6) | 6 (15.8) | 12 (15.4) | 0.698 |
| Heart failure composite | 0 (0) | 4 (4.7) | 0 (0) | 5 (6.4) | 0.251 |
| Atrial fibrillation | 0 (0) | 11 (12.8) | 4 (10.5) | 9 (11.5) | 0.264 |
| Stroke | 0 (0) | 4 (4.7) | 1 (2.6) | 6 (7.7) | 0.391 |
| All-cause mortality | 0 (0) | 5 (5.8) | 1 (2.6) | 15 (19.2) | 0.001 |
| Overall composite | 2 (5.7) | 24 (27.9) | 11 (28.9) | 40 (51.3) | <0.001 |

NYHA New York Heart Association; ApHCM apical hypertrophic cardiomyopathy; IL6 interleukin-6; CRP C-reactive protein; hsTnT high-sensitive Troponin T; NT-proBNP N-terminal Pro-B-type natriuretic peptide; LV left ventricular; EDV end-diastolic volume; ESV end-systolic volume; SV stroke volume; RV right ventricular; LA left atrial.

* Patients may have >1 events.
